# Step forward in epidemiological modeling: Introducing the indicator function to capture waves

**DOI:** 10.1101/2022.02.08.22270673

**Authors:** Abdon Atangana, Seda İğret Araz

## Abstract

In the last century mathematical modelled have been used to depict the dynamic spread of infectious. The aim is to determine the total numbers of infectious, recovered and susceptible classes, however the classes only represent accumulative. Having these classes one will be unable to predict waves, to determine a day to day numbers of newly infected cases some additional calculations are required. Collected data from real world situations are represented as day to day newly infections, from these data help to determine numbers of waves. The current existing mathematical models cannot be used to predict waves, while having known the predicted numbers of waves one can take several measures to control the situation. To solve this problem, we questioned the fact that the rates of infection and recovery are constant and suggested an indicator function, the function is obtained using experimental data. The indicator function is used as rate and introduced into the mathematical model. We considered a simple SIR model with different types of differential operators including classical and fractional derivatives. The models were solved numerically using well-known numerical schemes. The numerical solutions were plotted for different theoretical parameters, the obtained results depict real-world behaviours. To test the efficiency of this new approach, we collected data from South Afrıca and compared with experimental data. Not only our model waves but also it fit experimental data very accurately. This new approach will open new doors of investigation toward revolution to epidemiological modelling.

## 1 Introduction

Researchers use modeling to replicate real world problems and attempt to give a future behavior of such problems. This is usually achieved via mathematical models. Indeed to have very good predictions, data need to be analyzed, then the mathematical model is compared with collected data free of uncertainties. However, one of the most difficult part of this assignment is the conversion from observation to mathematical equations. Indeed if the conversion from observed facts to mathematical model is erroneous, the output will also lead to wrong prediction. To better understand the spread of a given infectious disease, mathematicians use mathematical models to replicate observed facts. In the last past years researchers have put much effort into modeling the spread of some infectious diseases[3]. However, when comparing collected data with mathematical models, there is most of the time a deviation from collected data and mathematical models, a clear indication that either the mathematics is erroneous or the data are uncertain. However, most of the time data are correct but there is a clear crossover behavior that cannot be captured by mathematical models. In particular, in this context, a concept known as rate of infectious or recovery or death is used in the mathematical model. However, in many published papers, this rate is considered as a constant, despite the fact that collected data show a clear indication that the rate is not constant; it is still assumed that the rate is a fixed proportion. Another point to raise is the plot represented by mathematical models, most of the time from such plots or predictions, it could be difficult to predict either the spread will have waves or not, in general, the prediction of accumulation values are presented, while a prediction of a day-to-day prediction is not known. This sometimes makes mathematical models results doubtful. Can we really use these models to predict future behavior of the spread? One of the most successful concepts used in this field is the reproductive number[5], but again so far the reproductive number may not be able to indicate to us either or not a spread of a given infectious disease will lead to multi-waves. To properly achieve the conversion from observed facts into mathematical formulas, there are few concepts that should be revised.

1. What is the rate of infection?
2. What is the strength of the infections?
3. What are the crossover patterns presented by the spread?
4. How can day-to-day new infections numbers be predicted?
5. What differential operator should be utilized to model a given spread and how to identify it?

In this article, some of the above points will be investigated with the aim to move the field of epidemiological forward.

## 2 Existing theories

One of the greatest contribution made by mathematicians in the field of biological modeling is perhaps to construct mathematics models able to replicate dynamic spread of infectious disease. Indeed several theories have been suggested in the last past years and have been used to give advice regarding the spread of infectious diseases. For example, the SIR model was suggested to predict the accumulative number of Susceptible (S), Infectious (I) and Recovery (R) classes[4]. This model is obtained using some important parameters including the rate of infection, the rate of recovery. In the last decades, these rates have been considered to be constant, however when collecting data from real-world situation it shows that the rate maybe a function of time. More importantly, the concept of reproductive number has been introduced to be an indication of the stability of the model and guide regarding the survival or the extinction of the virus. Indeed, some predictions have been accurately done, however, these theories did not help indicate either or not the spread will have waves. This is very important for decisions making, if the mathematical theories or models can indicate either or not the spread will have waves, measures could be taking by medical and governmental authorities on how the spread could be contain to avoid waves.

In the general to recap, we consider the following SIR model

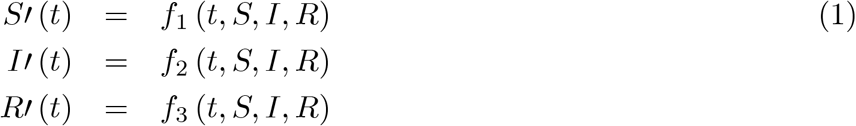

The above system is always subjected to initial condition *S*(0), *I*(0) and *R* (0) these are all positive or zero. The analysis always start off with the proof of positive solutions, where it is requested to show that for any time, the solutions *S*(*t*), *I* (*t*) and *R*(*t*) are positives. The second step is to establish the equilibrium points that will be later used to formulate the reproductive. The Lyapunov analysis can be followed, the local and global stability of the models can be established.

As explained in the paper by Atangana and Araz [6], the detection of waves can be obtained following steps.

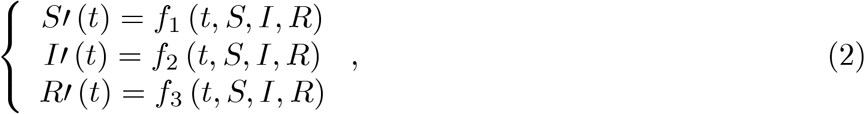

we solve equations

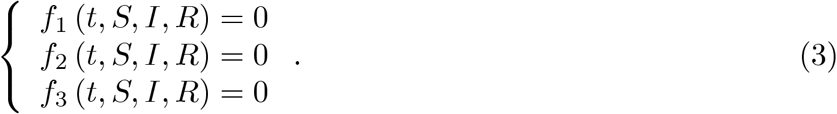

(*S*\* *I*\* *R*\*) are obtained, then

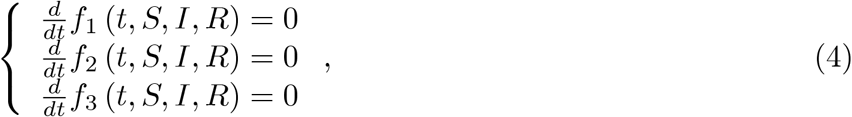

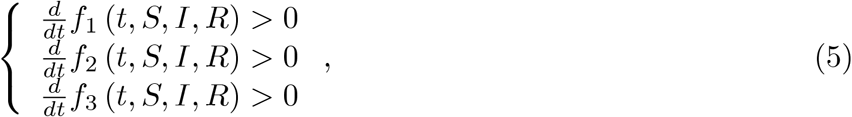

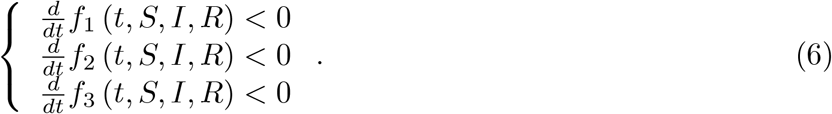

The conditions under which the above are satisfied can be obtained. Using the second derivative analysis, concave up and concave down can be detected.

## 3 New approach: Introduction of a daily indicator function

In order to take into account real changes occurring in real world scenario, we introduce in this section an alternative way to obtain an epidemiological model with constant rates. Let us consider the following simple SIR model:

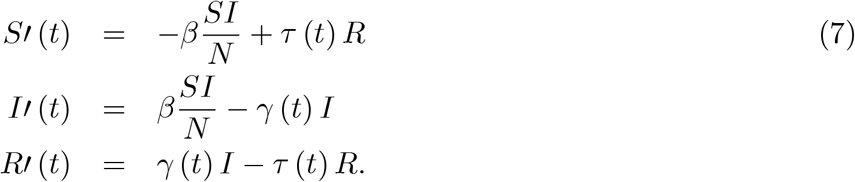

Here we assume that *γ* = *γ* (*t*), *τ* = *τ* (*t*) are the functions of time that are obtained using experimental data as: For example, after collection of new daily infection (*d*_1_,*d*_2_,..,*d*_*n*_)

The rate between two days can be calculated as

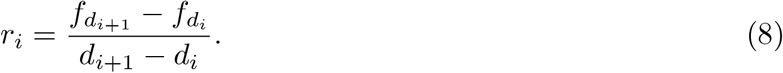

where 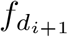 is the observed data of day *d*_*i*+1_ The daily indicator function can be obtained as

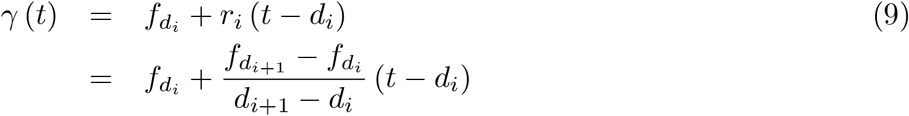

and

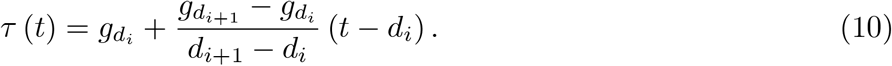

Thus, the model can be reformulated as:

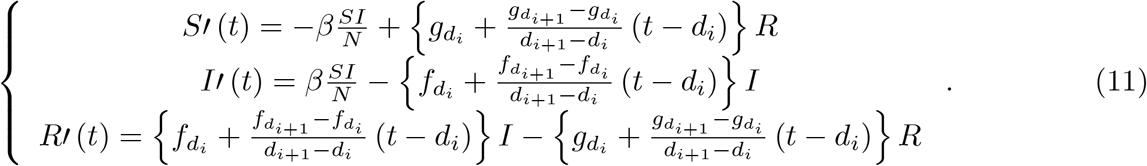

The above model can be used to depict more accurately with waves the spread from *d*_1_ to *d*_*n*_. However, to obtain a suitable prediction, we need to interpolate by obtaining *γ*_max_(*t*), *γ* (*t*) and *γ*_min_ (*t*). These functions can be obtained using moving average of statistics.

The model becomes

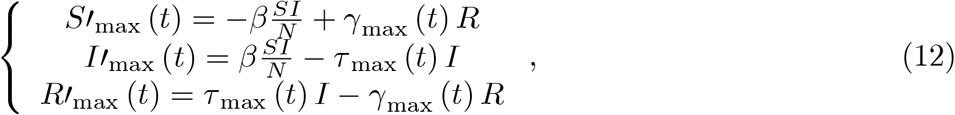

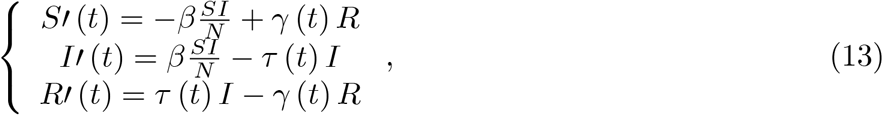

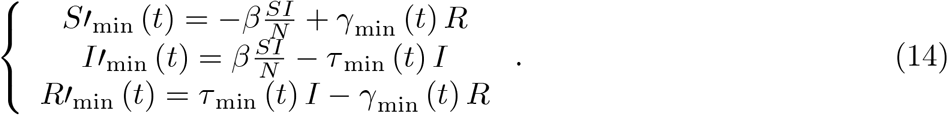

Noting that *γ*_max_ (*t*) = *γ* (*t*) = *γ*_min_ (*t*) for *t* ∈ [*d*_1_,…, *d*_*n*_]

The above system can be solved using any accurate numerical scheme for example Adams-Bashforth for classical system we get

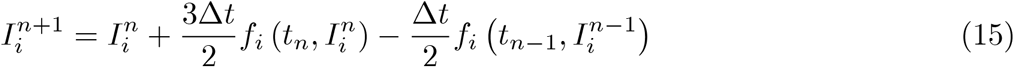

where *I*_1_ = *S, I*_2_ = *I* and *I*_3_ = *R*. To test the effectiveness of the methodology, we present here some numerical simulations presented. Additionally, collected data from South Africa are considered and compared with the suggested model. The data are collected from 5 March 2020 to 28 January 2022.

**Figure 1.**
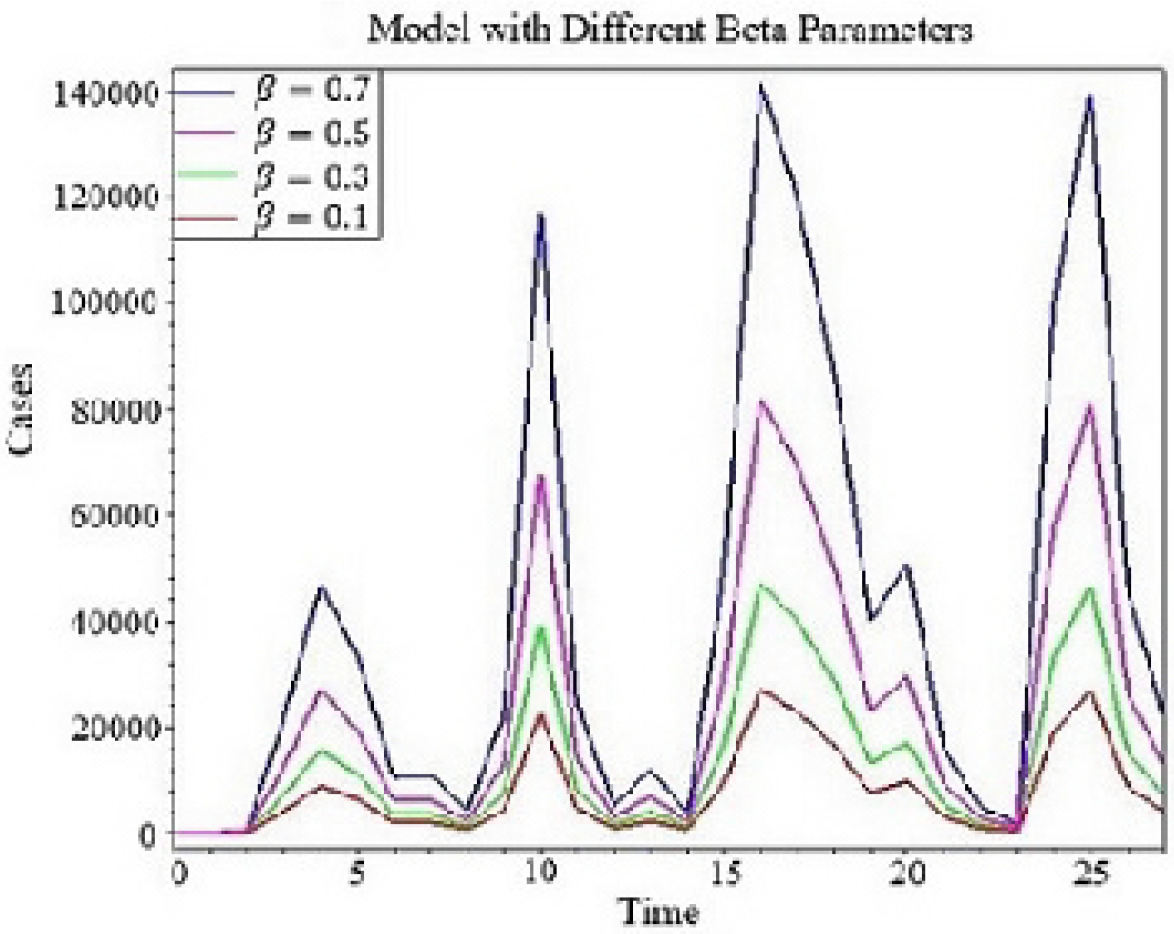
Numerical simulation of model with different *β* parameters.

**Figure 2.**
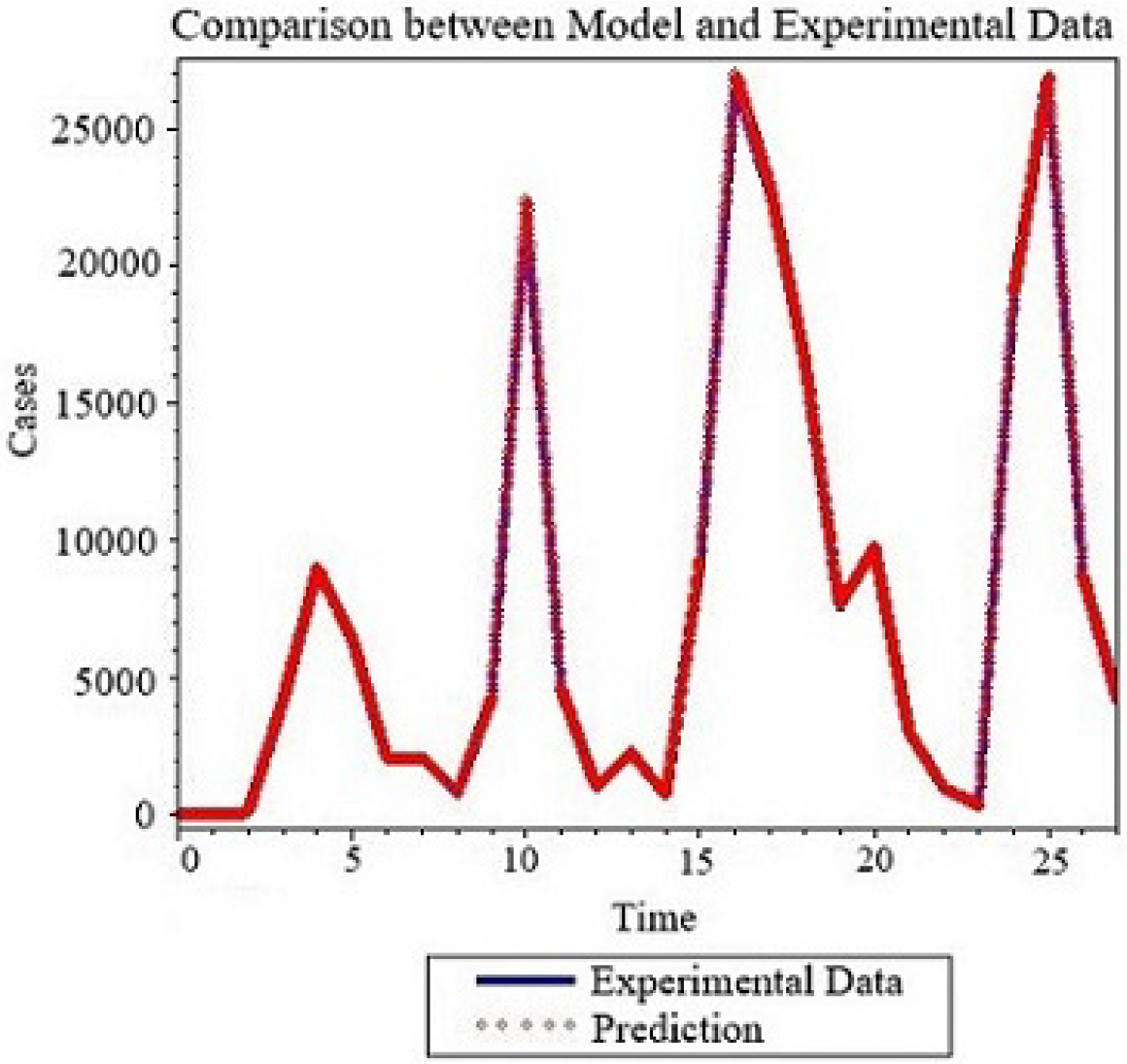
Numerical comparison between model with classical derivative and experimental data for *β*= 0. 1.

### 3.1 Model with Caputo-Fabrizio derivative

The model presented above is clearly depicting process or spread with classical behaviors only, such behaviors can be studied using the derivative based on the rate of change as we presented above. Nevertheless, it also observed in several real world problems classical behaviors convoluted with fading memory patterns[1]. This class of real world problems has been recognized to be efficiently modeled using the Caputo-Fabrizio type of differential operator. Therefore, in order to include into the mathematical model the effect of fading memory, we replace the classical differentiation with Caputo-Fabrizio to obtain

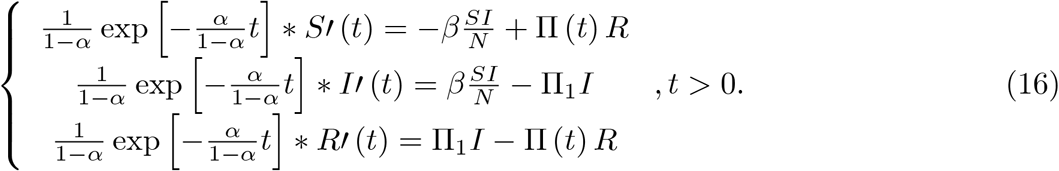

where

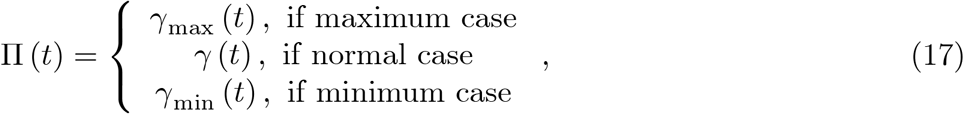

and

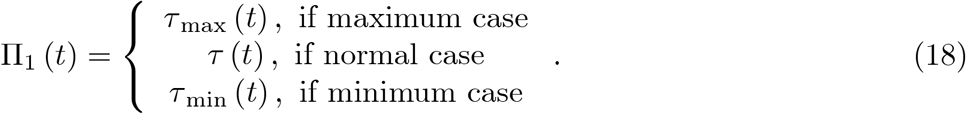

The initial conditions should be

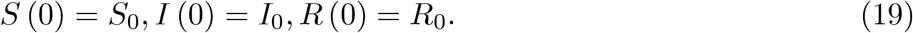

Of course, the solution of the above system may not satisfy the initial conditions since they are clearly processes with memory. It is also not practical to replace the parameter *β* with a specific power to justify the dimensionalization. These parameters are data from real world situation therefore should not be modified for any reason. The system can also be solved using an accurate numerical scheme for example

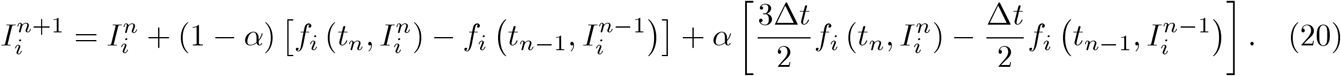

Numerical simulations are presented below for different values alpha.

**Figure 3.**
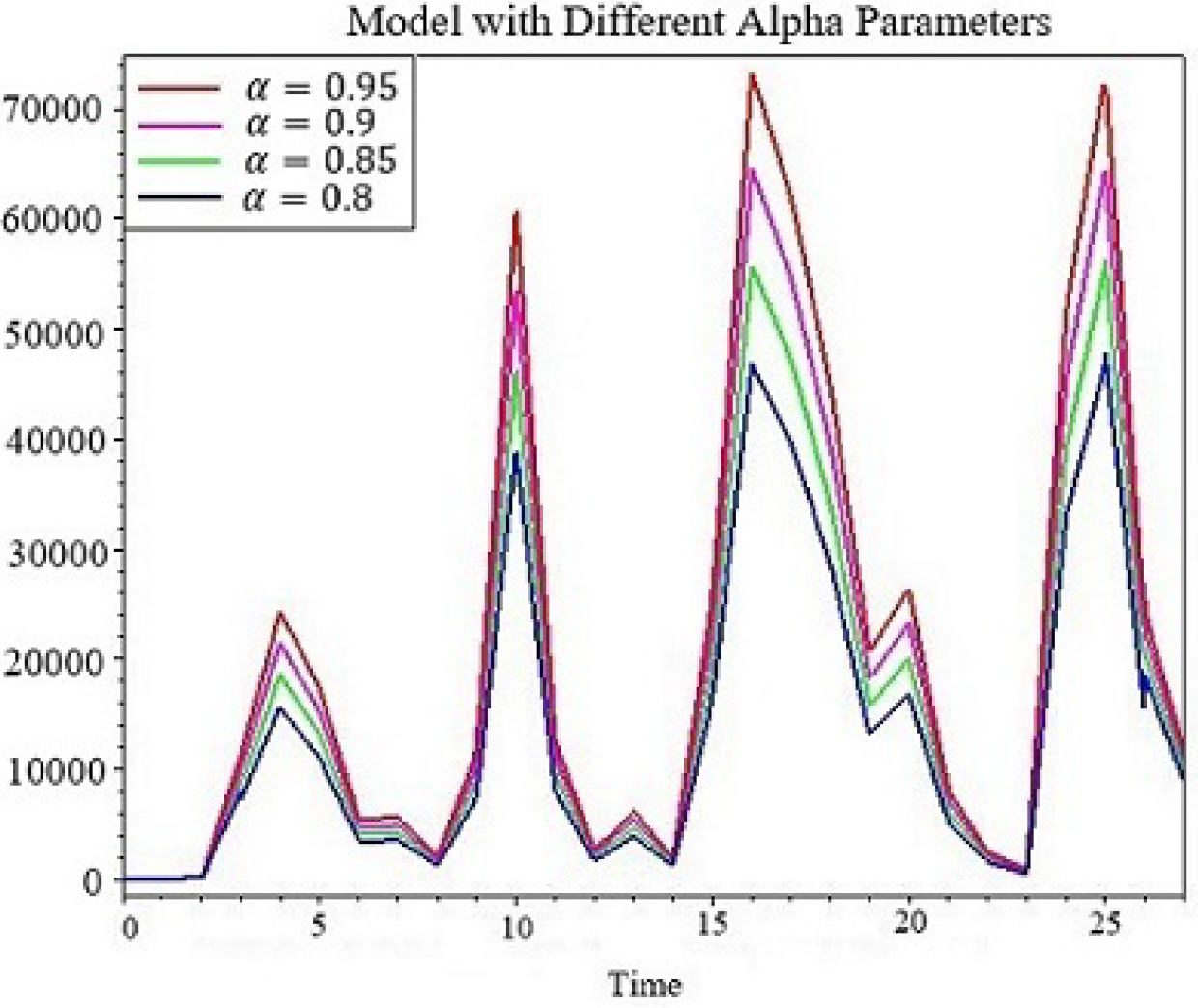
Numerical simulation of model with Caputo-Fabrizio derivative for different values of *α, β* = 0.5

**Figure 4.**
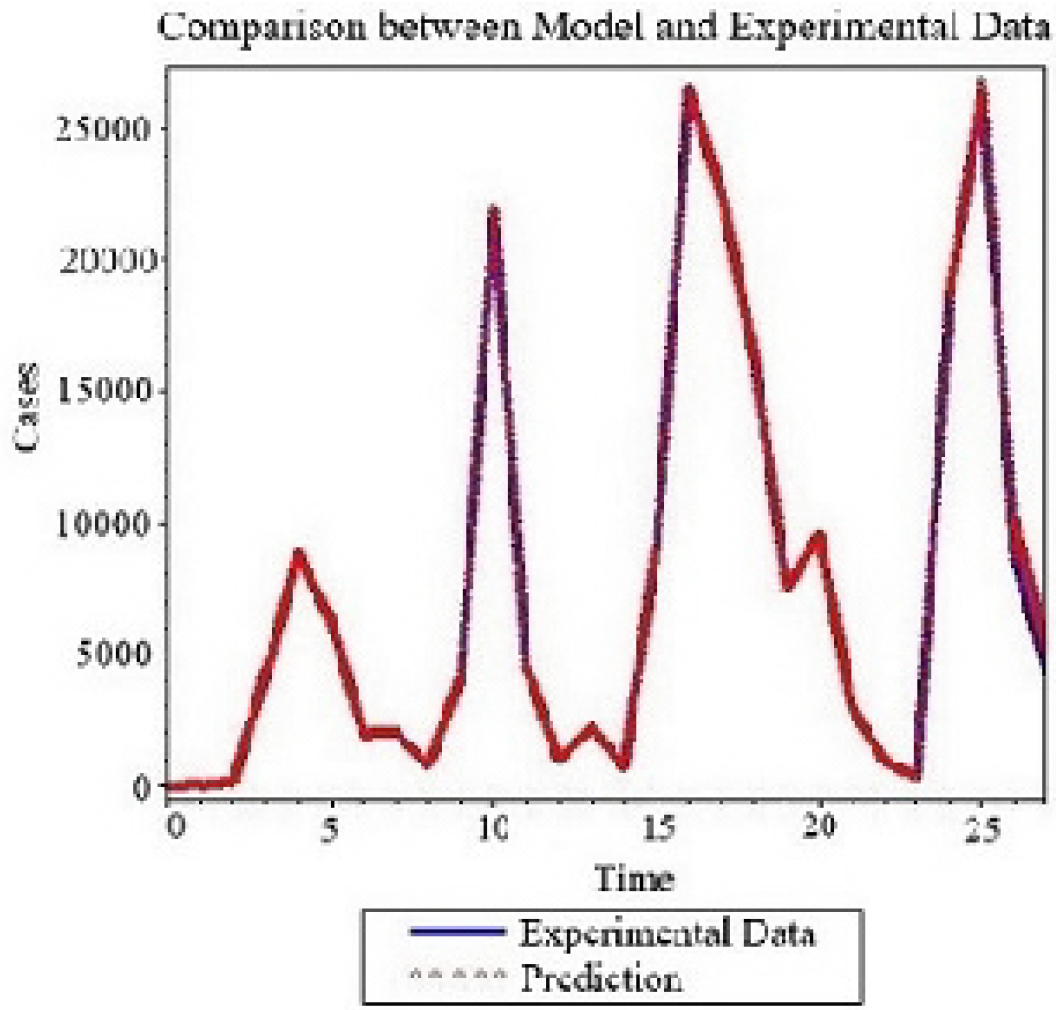
Numerical comparison between model with Caputo-Fabrizio derivative and experimental data for *α* = 0.8, *β* = 0.6

**Figure 5.**
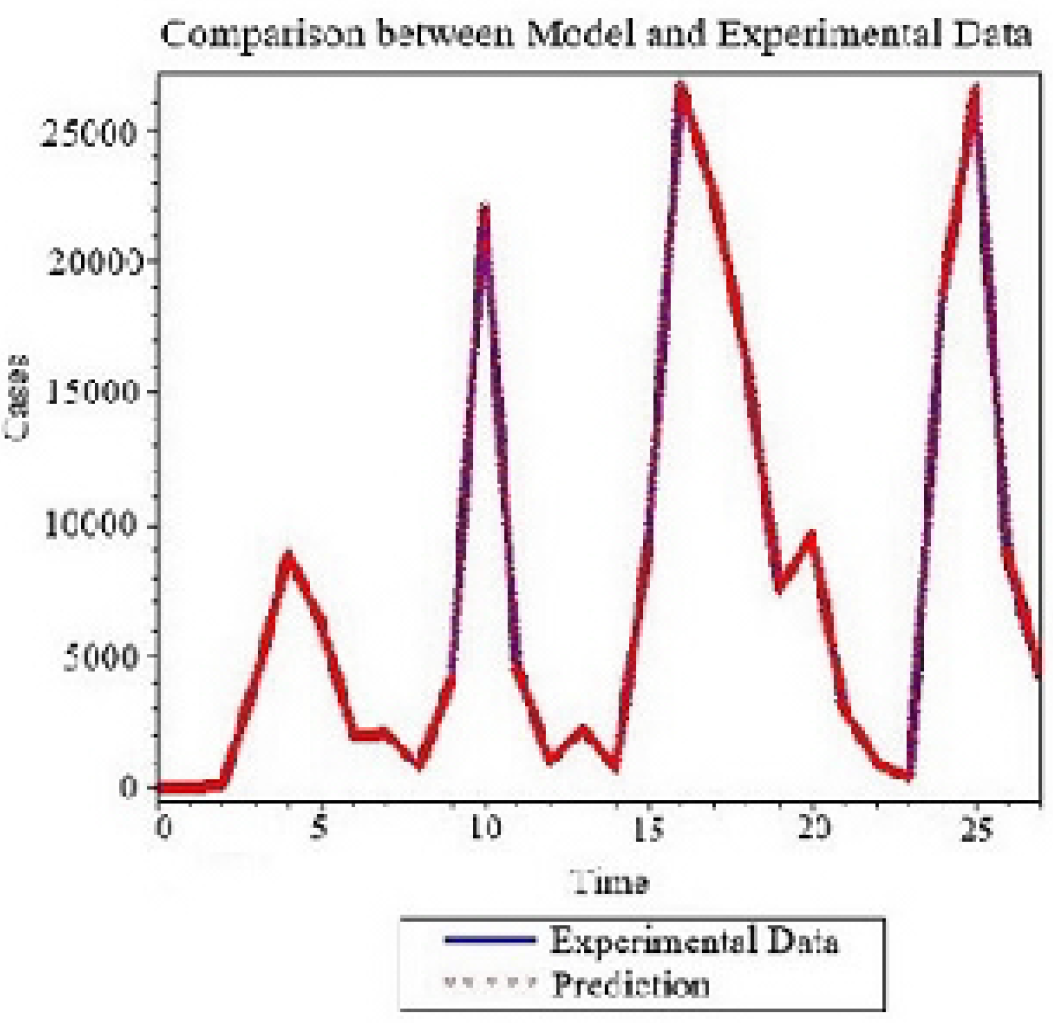
Numerical comparison between model with Caputo-Fabrizio derivative and experimental data for *α* = 0 .9, *β* = 0.23.

**Figure 6.**
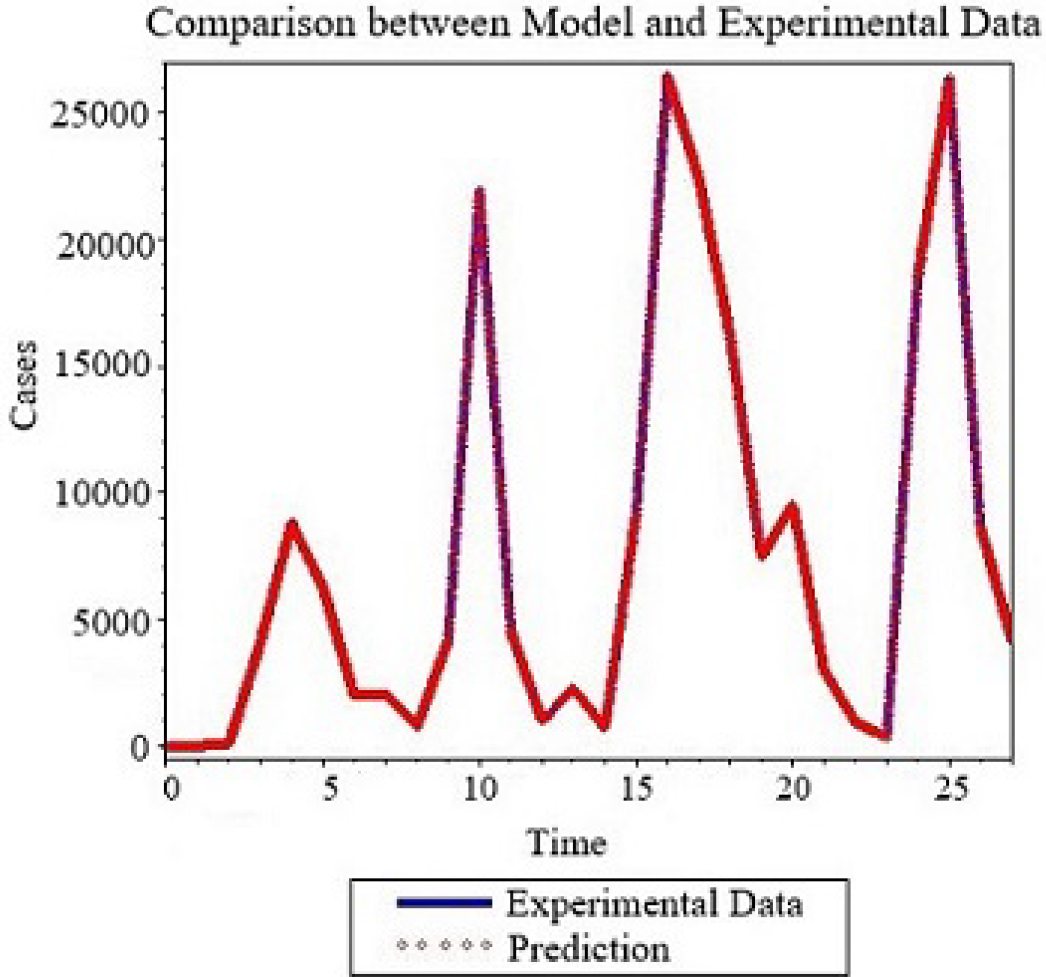
Numerical comparison between model with Caputo-Fabrizio derivative and experimental data for *α* = 0.95, *β* = 0.16.

**Figure 7.**
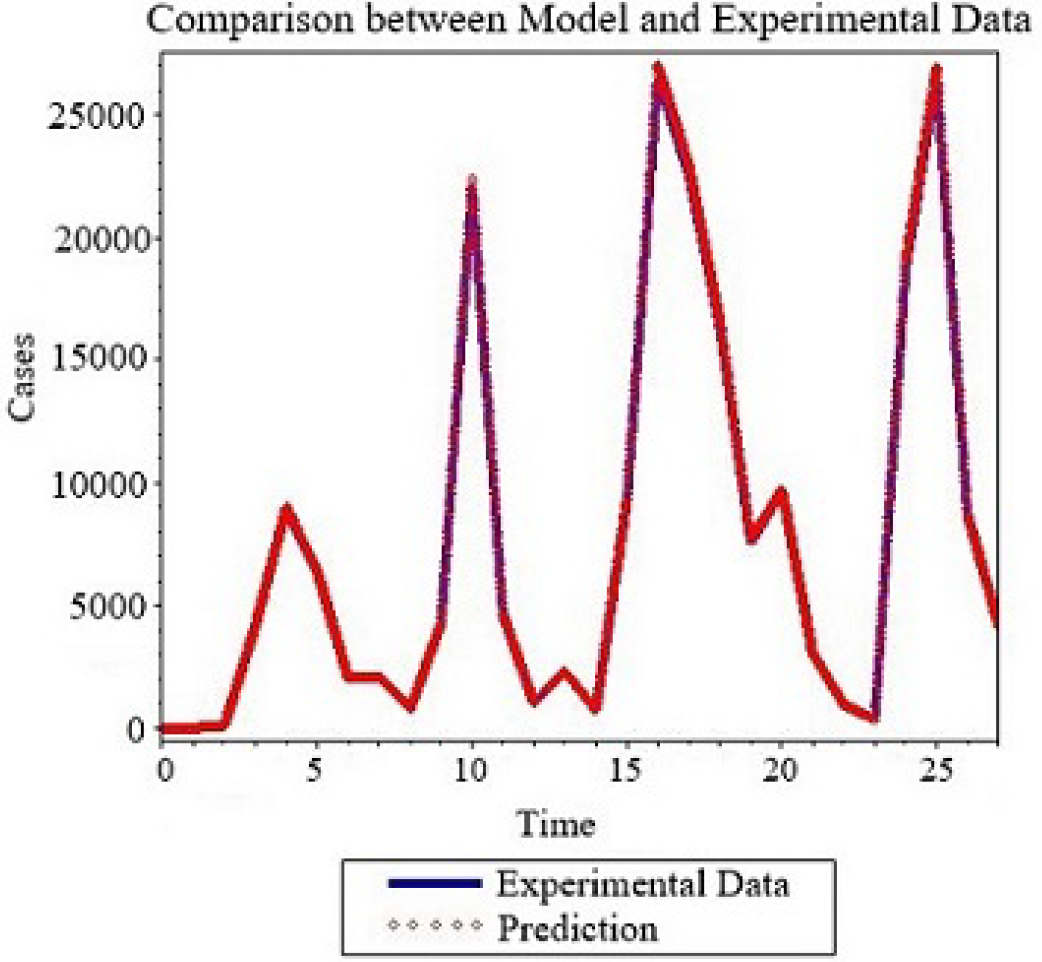
Numerical comparison between model with Caputo-Fabrizio derivative and experimental data for *α* = 1, *β* = 0.1.

### 3.2 Model with Caputo derivative

In the same line of idea, we have some spread exhibitng changes convoluted with power-law behaviors. These behaviors are sometimes characterized by long range or non-Gaussian behaviors. Since these behaviors occurred as the function of time, we therefore replace the classical differentiation by the Caputo fractional derivative

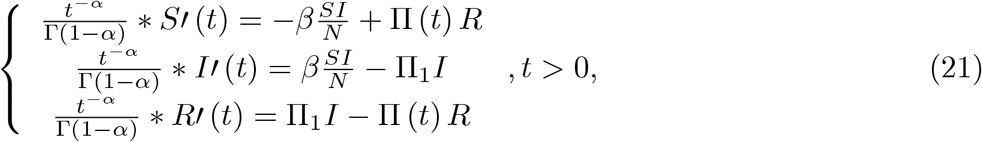

Then the numerical scheme of system can be obtained as follows

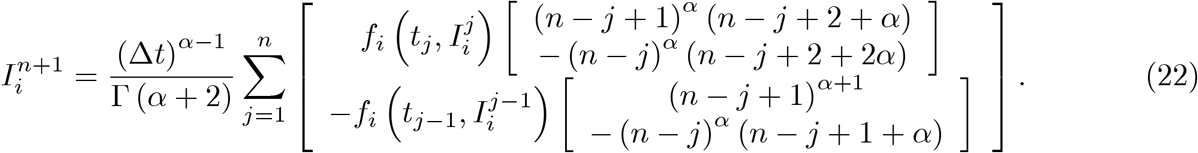

**Figure 8.**
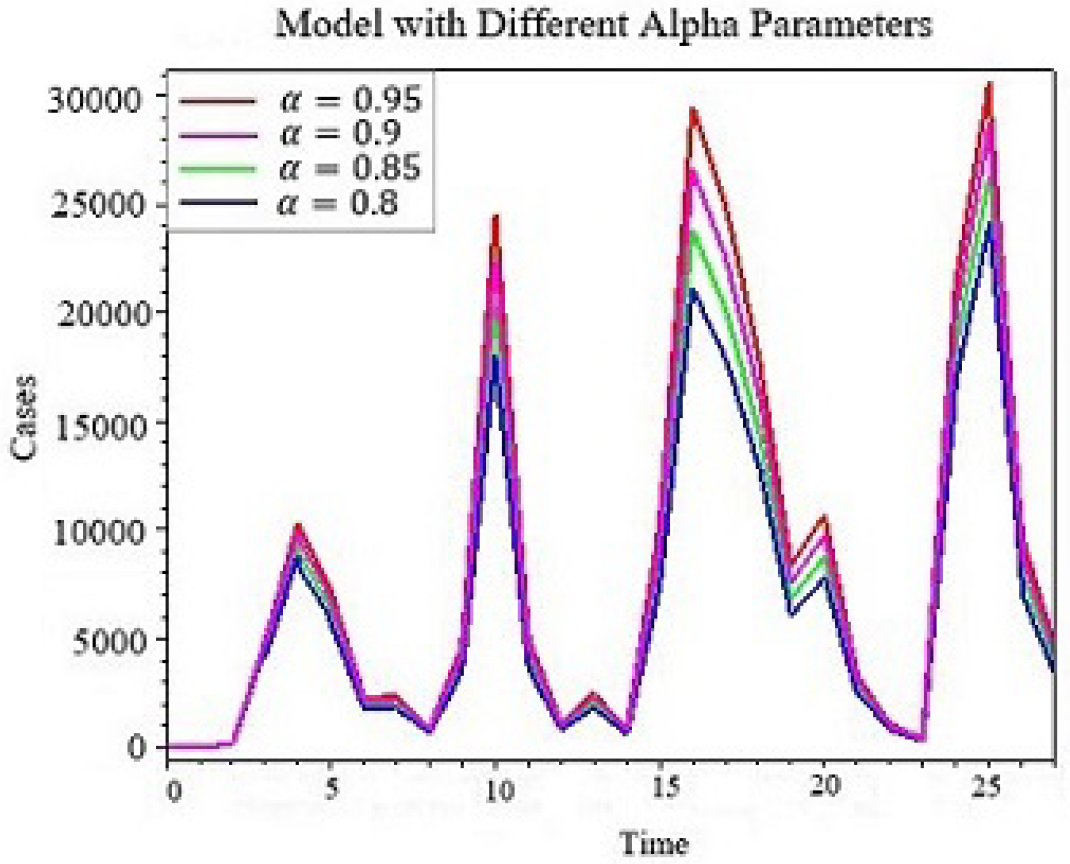
Numerical simulation of model with Caputo derivative for different values of *α, β* = 0. 5.

**Figure 9.**
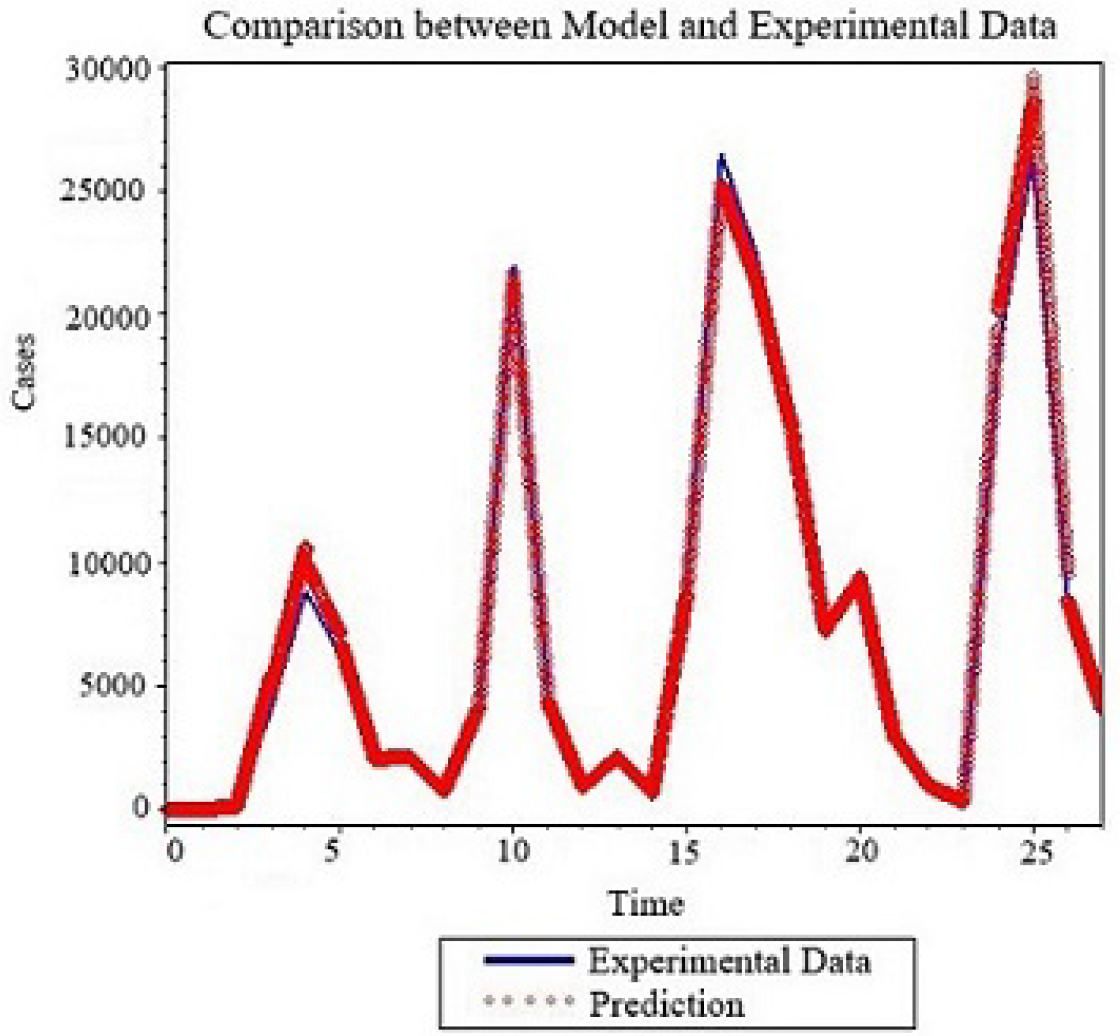
Numerical comparison between model with Caputo derivative and experimental data for *α* = 0.8,*β* = 33.

**Figure 10.**
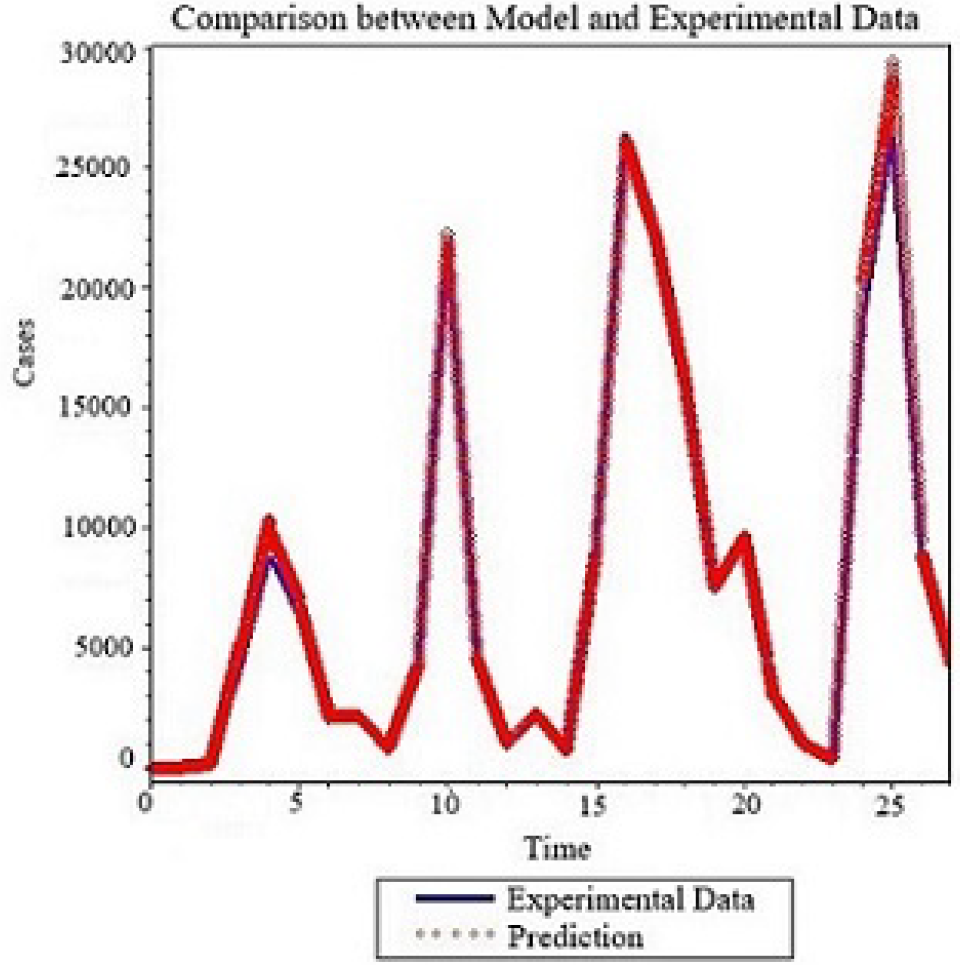
Numerical comparison between model with Caputo derivative and experimental data for *α* = 0 .85, *β* = 31.5.

**Figure 11.**
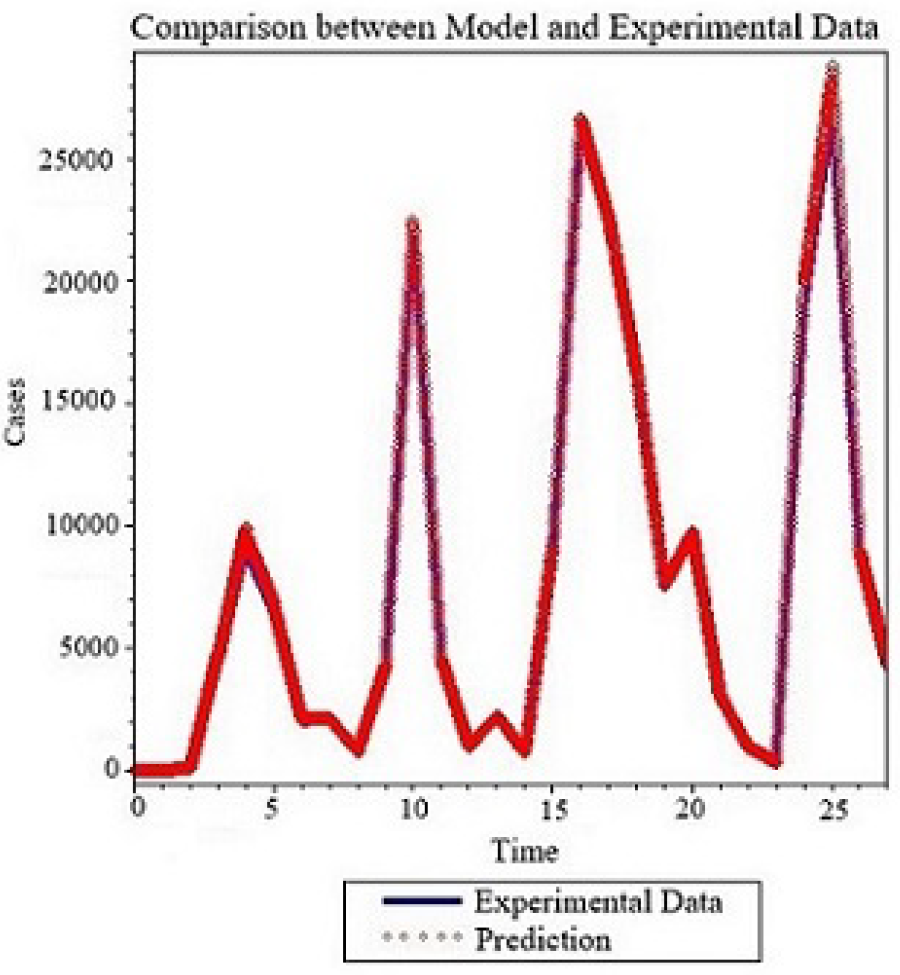
Numerical comparison between model with Caputo derivative and experimental data for *α*= 0.9, *β* = 30.

**Figure 12.**
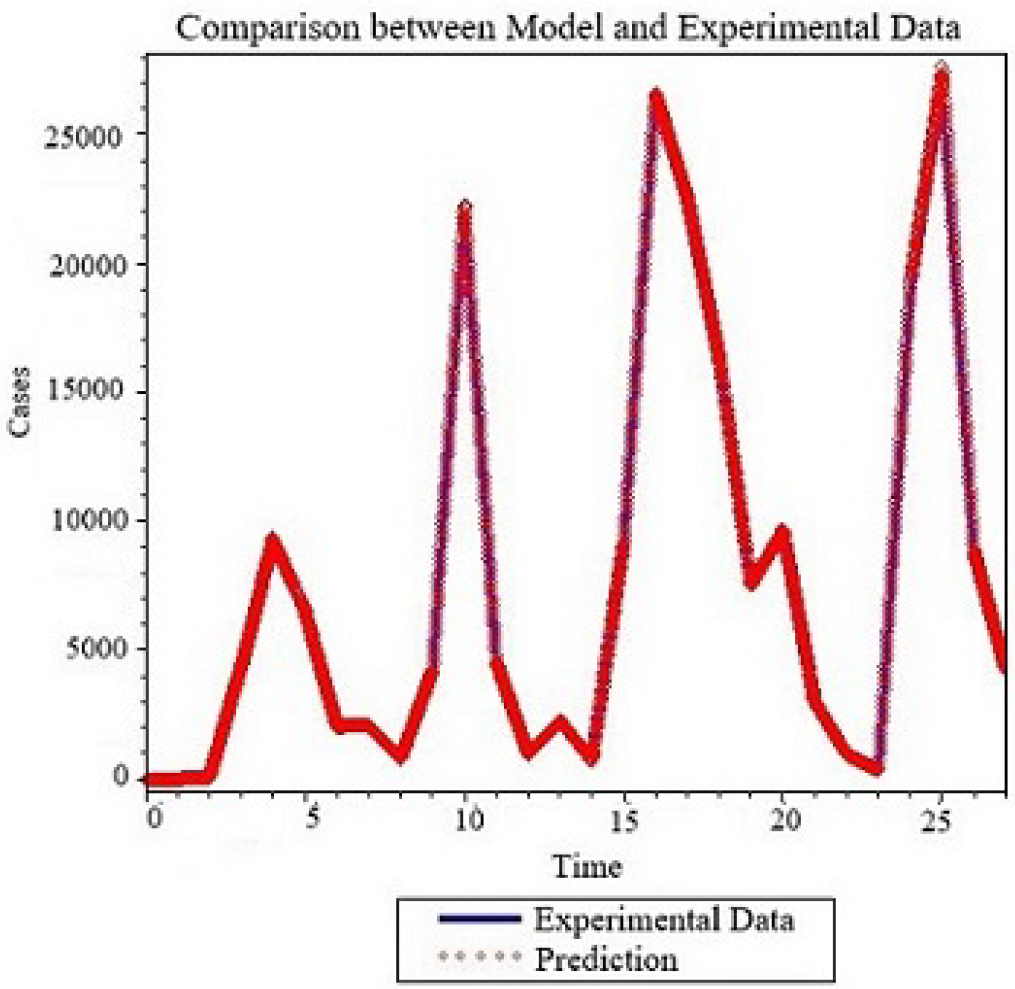
Numerical comparison between model with Caputo derivative and experimental data for *α* = 0.95, *β* = 28.5.

### 3.3. Model with Atangana-Baleanu derivative

To include into the crossover from fading to power law, the rate of change will be convoluted with the generalized Mittag-Leffler kernel[2]. The modified SIR model is presented as follow below. The modified model will be solved using some known numerical method. The obtained numerical solutions are depicted for several fractional orders. In addition, data collected from different countries are compared with the mathematical models.

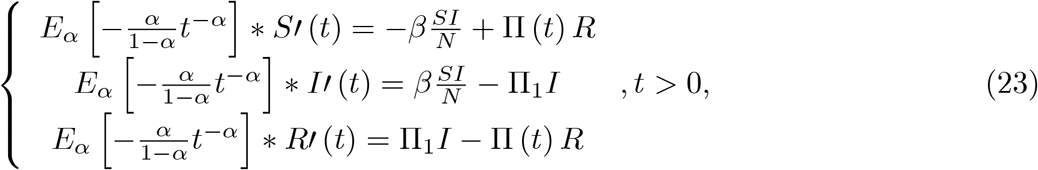

Then the system can be solved numerically as follows;

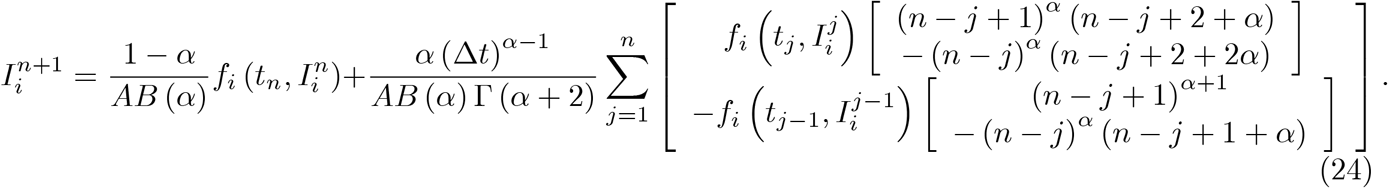

**Figure 13.**
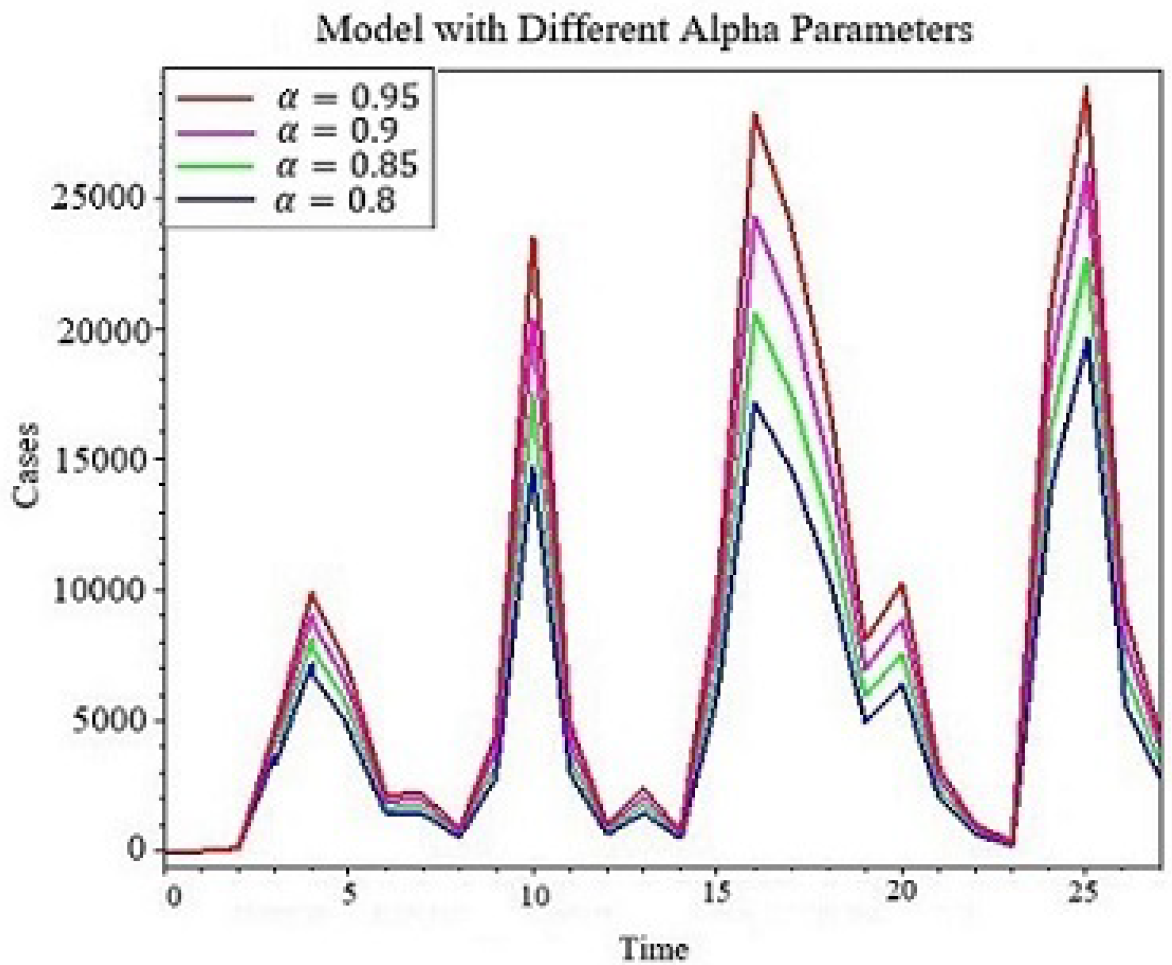
Numerical simulation of model with Atangana-Baleanu for different values of *α, β* = 30.

**Figure 14.**
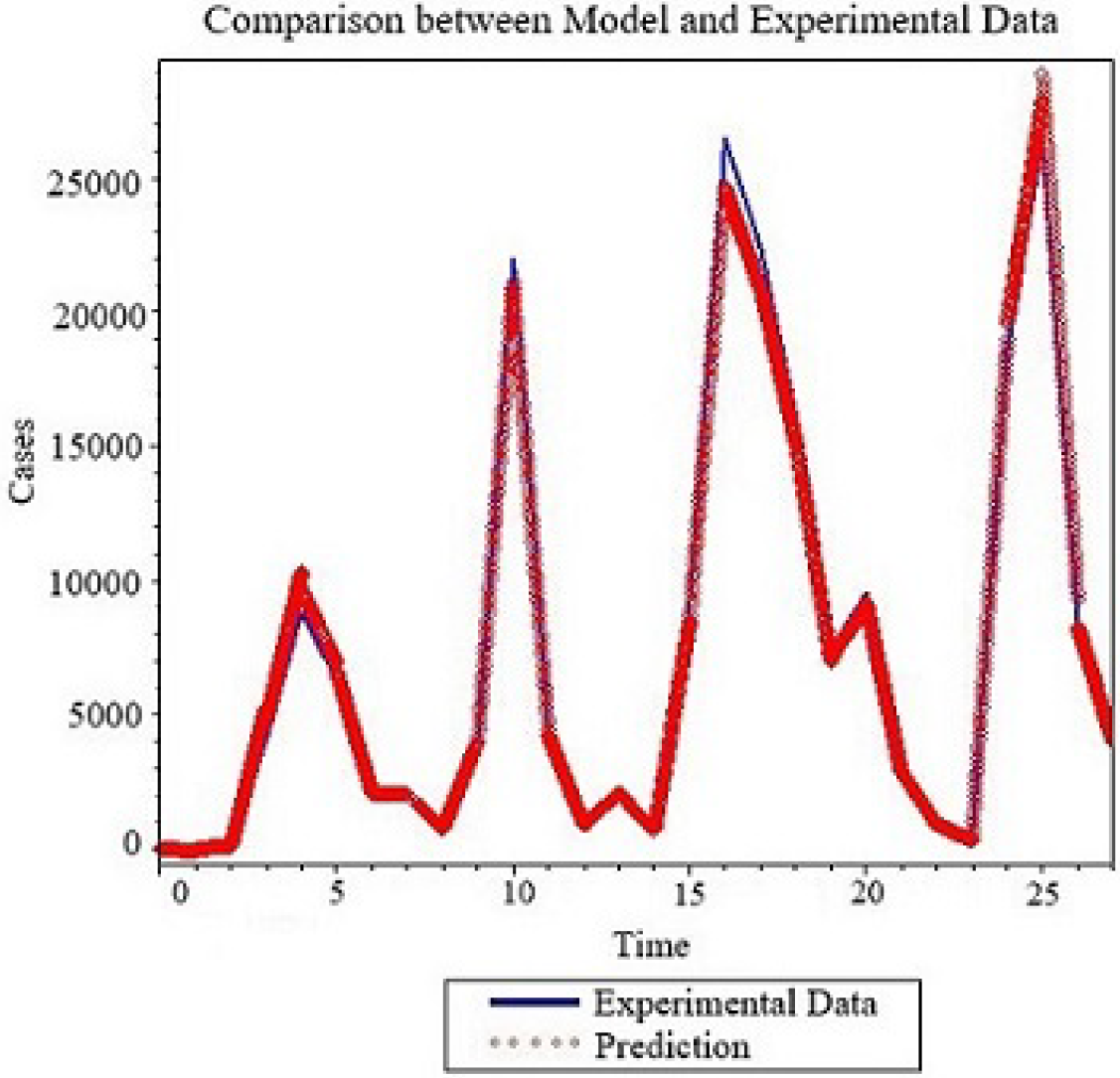
Numerical comparison between model with Atangana-Baleanu derivative and experimental data for *α*= 0.8, *β* = 36.

**Figure 15.**
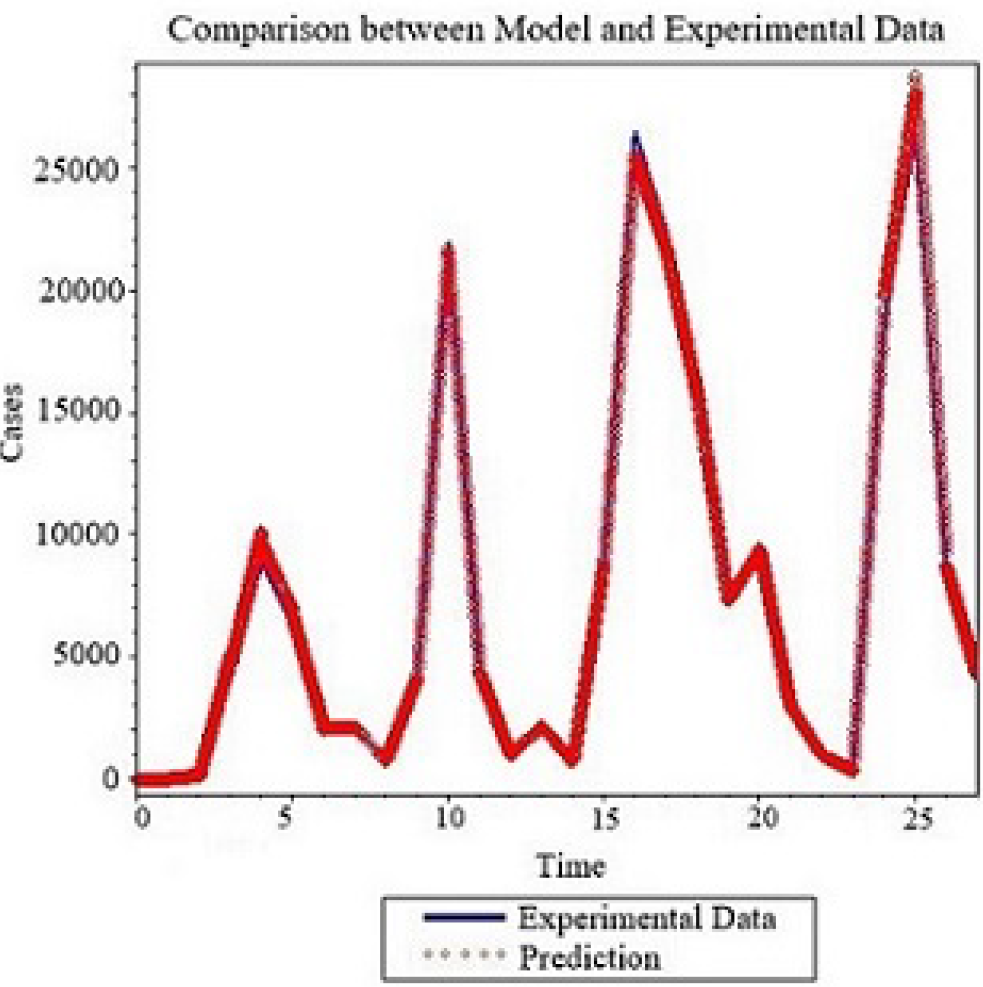
Numerical comparison between model with Atangana-Baleanu derivative and experimental data for *α* = 0.85, *β* = 33.

**Figure 16.**
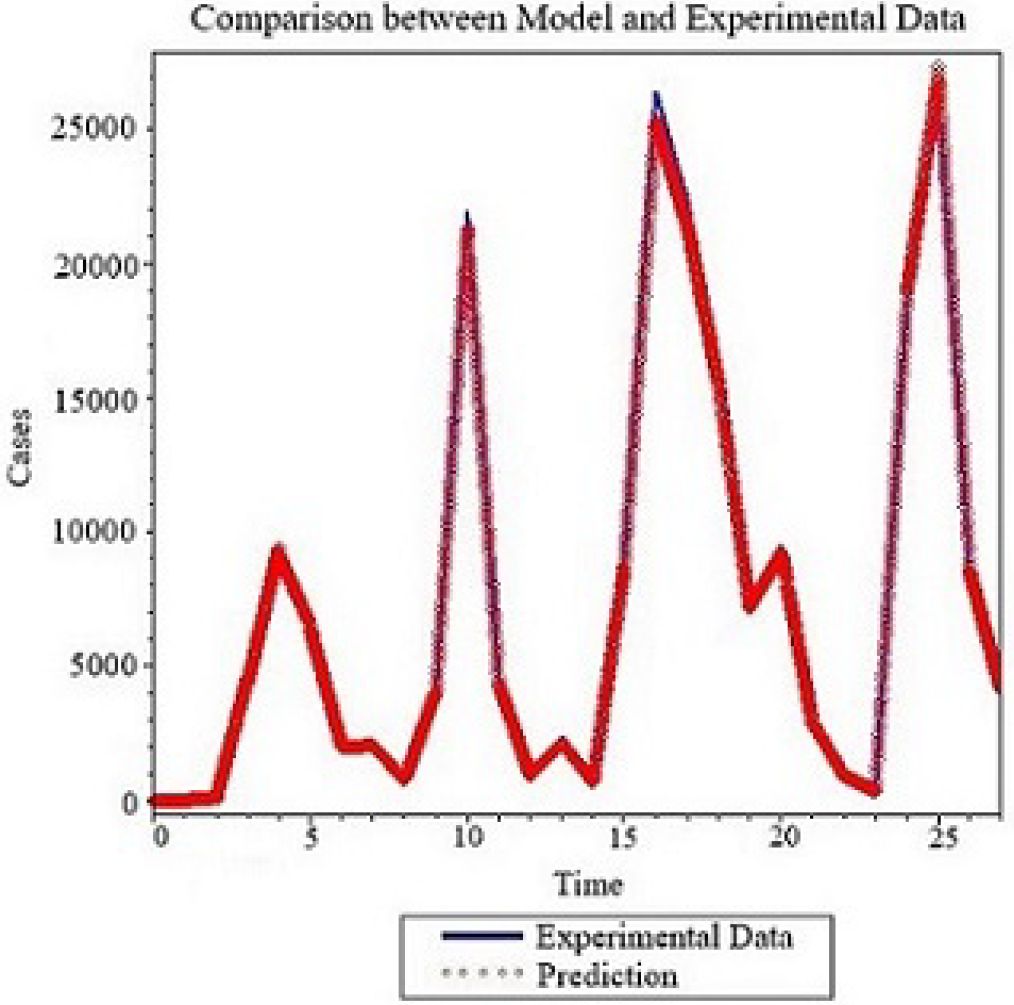
Numerical comparison between model with Atangana-Baleanu derivative and experimental data for *α* = 0.9, *β* = 30.

**Figure 17.**
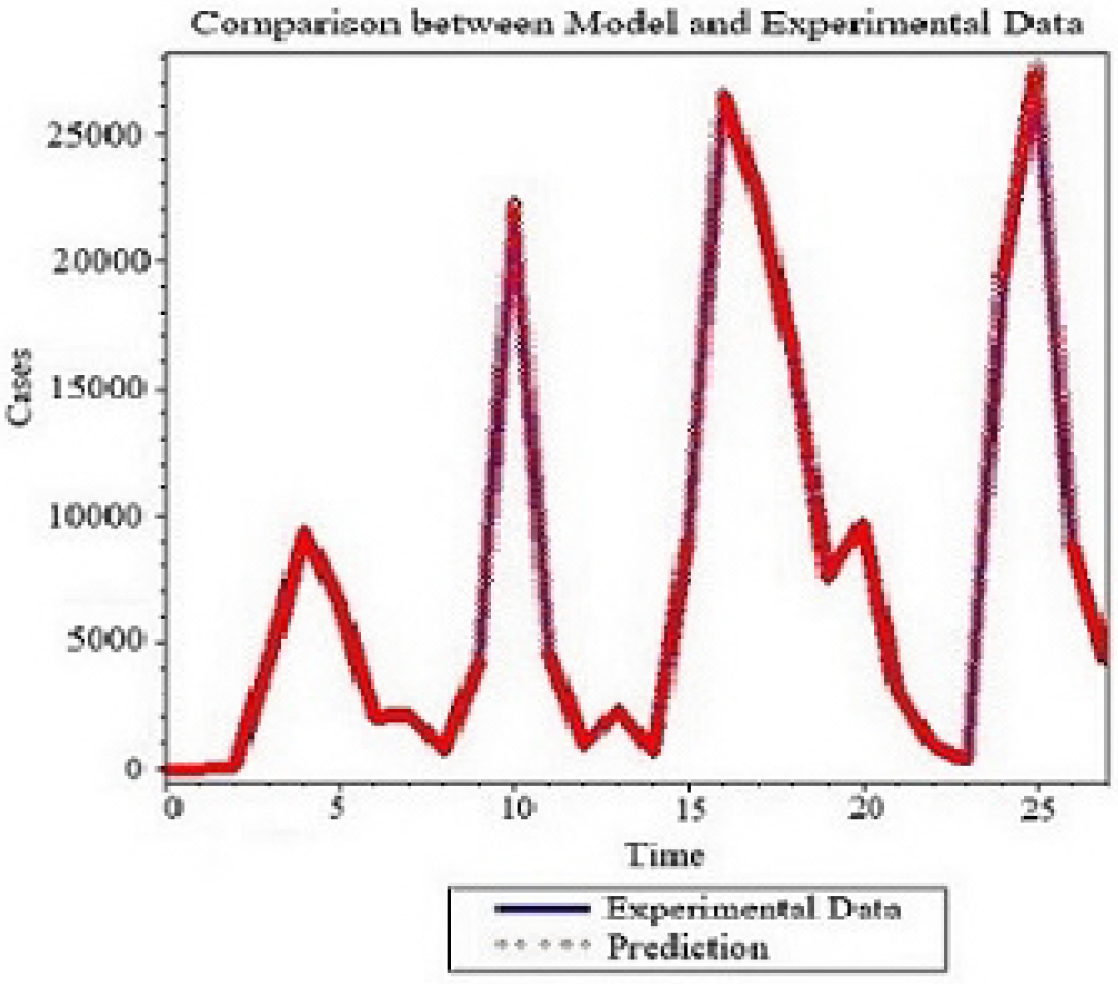
Numerical comparison between model with Atangana-Baleanu derivative and experimental data for *α* = 0.95, *β* = 28.5.

## 4 Prediction

In this section the suggested models are used to predict future behaviours of the spread. Data are collected from South Africa[7]. Four cases are considered, including the model with Caputo, Caputo-Fabrizio, classical and Atangana-Baleanu derivatives. For each model, three graphs are obtained for infected class, the first graph corresponds to the upper boundary that often describe the worst-case scenario, the second graph is the projection of the normal situation and then last graph represent the ideal scenario where less infection is predicted. The obtained results should that South Africa may soon face a fifth wave that could record more infection than the previous wave in the worst-case scenario. In normal and ideal scenario there is still a possibility of fifth wave but with less infections than the previous waves. If the worst-case scenario occurs, it could be due to mutation of the virus, for example the Omicron was more infectious than its predecessors. The numerical simulations are depicted in figure below for different models. We stress on the fact that our model did not include all factors therefore it could deviate from the real-world scenario, however under the same condition described here both countries will face fifth very soon. In the case of South Africa, this period correspond to winter period.

**Figure 18.**
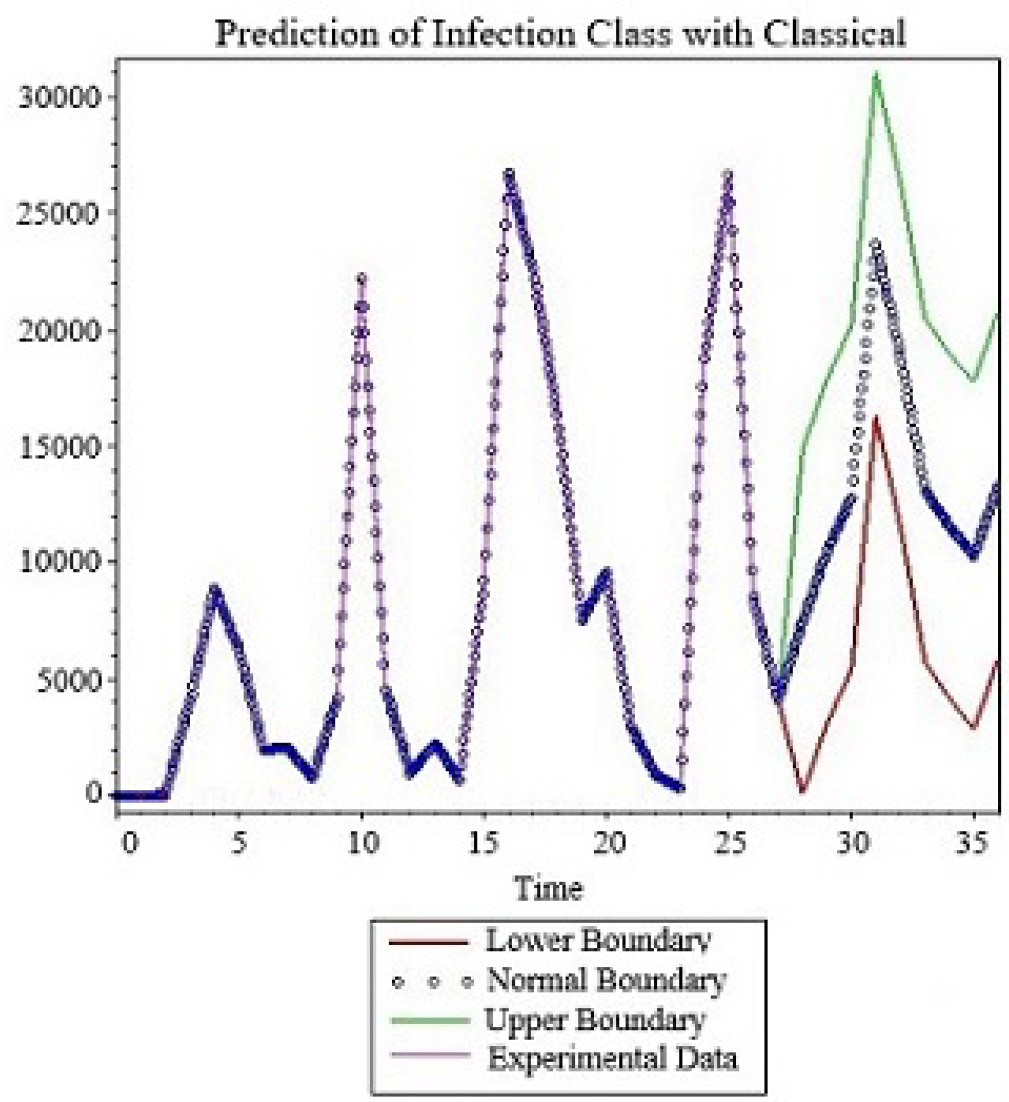
Numerical simulation for prediction of infection class with classical derivative for *β*= 0.07.

**Figure 19.**
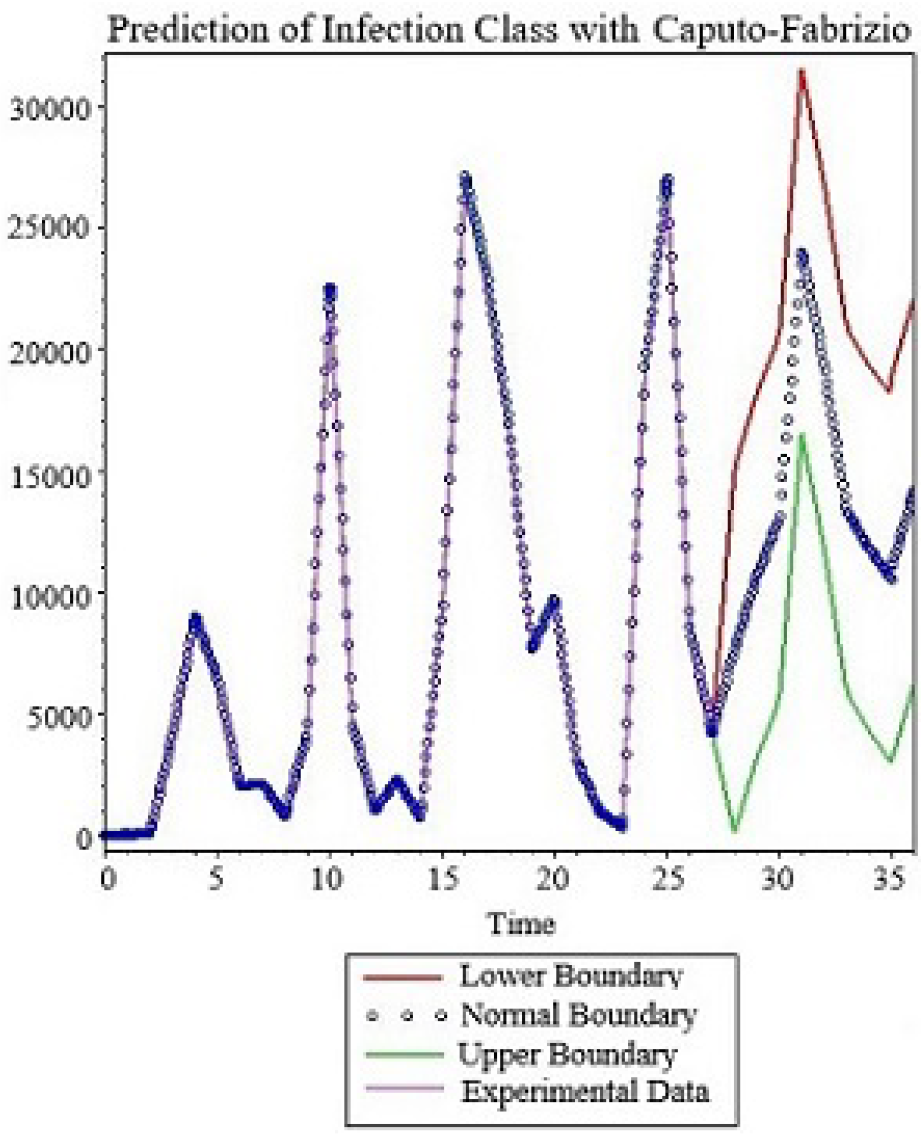
Numerical simulation for prediction of infection class with Caputo-Fabrizio derivative for *α* = 0.9, *β* = 0.23.

**Figure 20.**
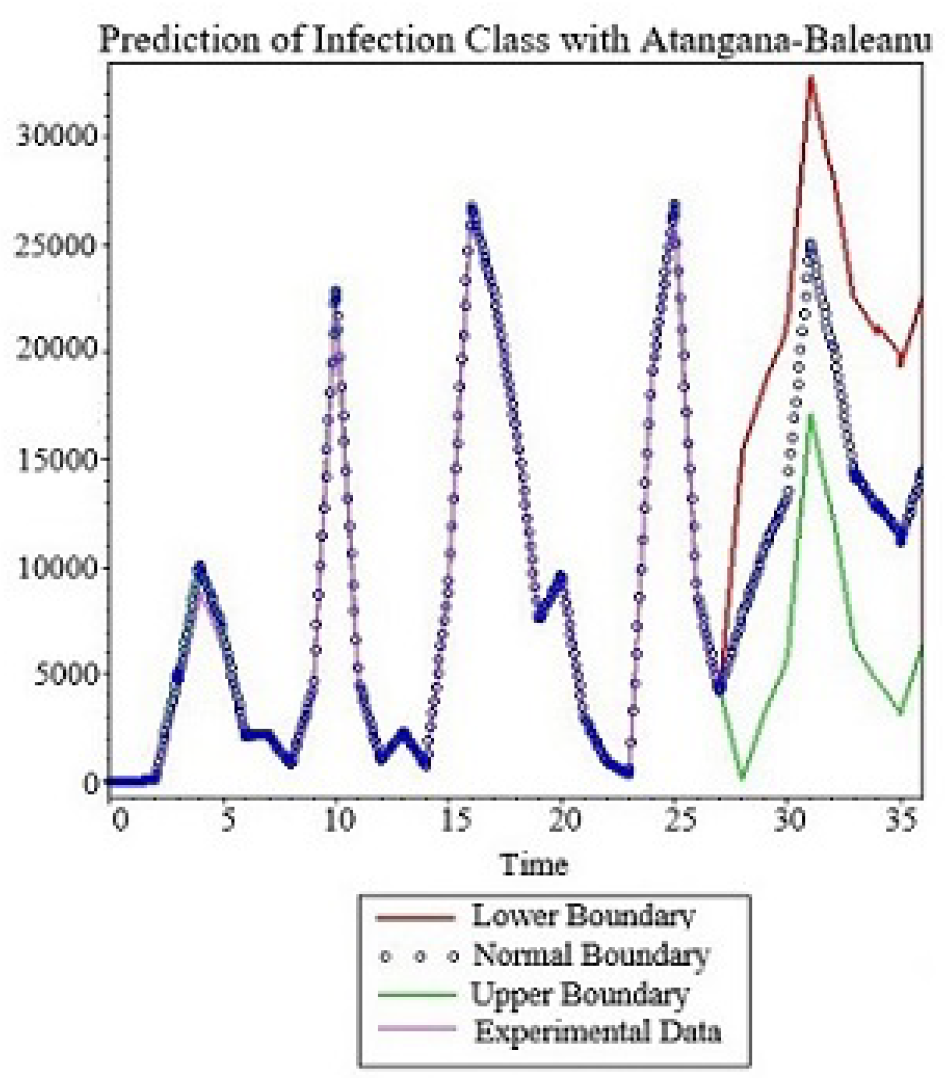
Numerical simulation for prediction of infection class with Atangana-Baleanu derivative for *α* = 0.9, *β* = 23.5.

**Figure 21.**
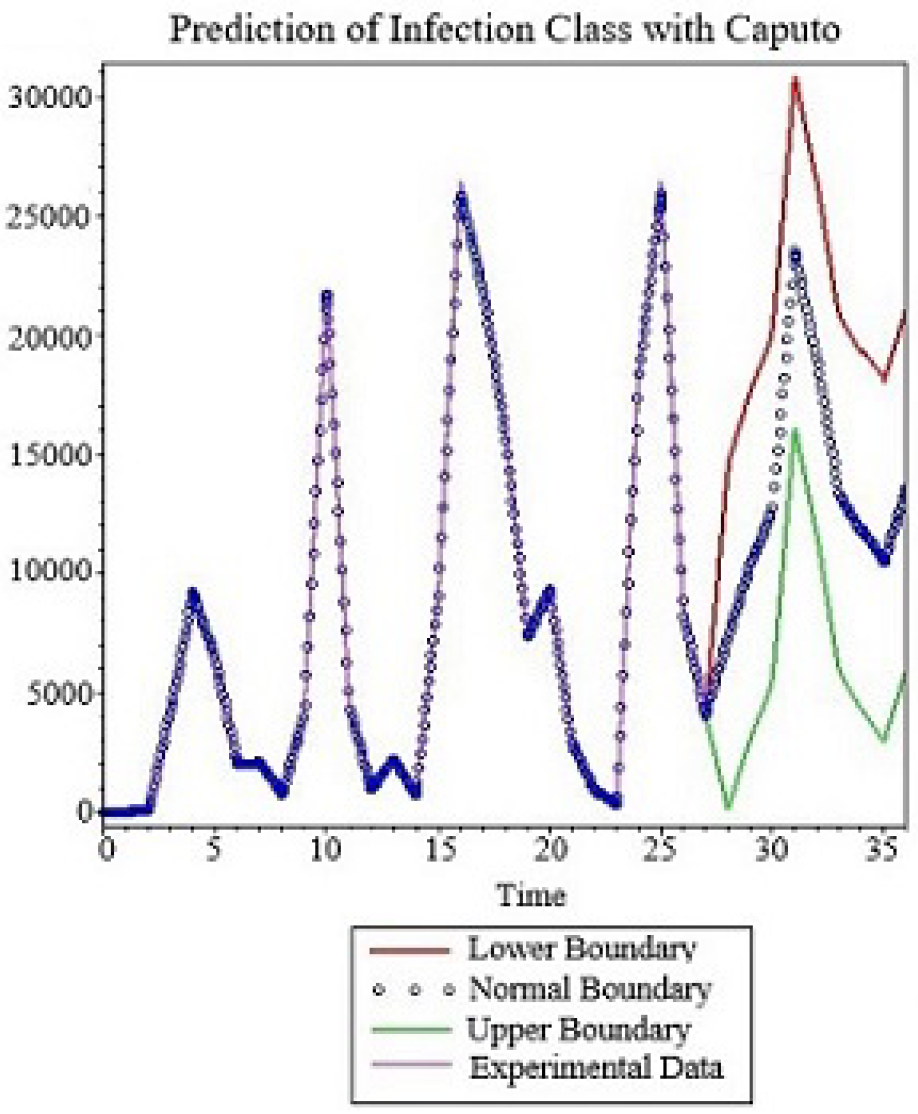
Numerical simulation for prediction of infection class with Caputo derivative for *α* = 0.95, *β* = 21.1.

## 5 Conclusion

Mathematical models are made to replicate dynamical process observed in nature. This should also be true for models used to depict spread of infectious diseases. Several advance studies have been done to make these models more accurate, for example the reproductive number was introduced to study the stability of the model and predict if the model will predict a raise in spread or decline of the spread. Several differential operators were used with aim to include into mathematical models’ complex behaviours of the spread, however, on several occasions, these mathematical models failed to not only predict the accumulative numbers for infectious classes but also, they failed to indicate either or not the spread will have waves. Failures to accurately predict the dynamic of the spread will lead automatically to wrong prediction thus, wrong decisions will be taken by law makers regarding the spread. Noting that any revolution in any field of study starts when existing theories are put in test, or when they do not replicate accurately. With no doubt existing models have failed in several instances, for example existing models have failed to predict waves for the Covid-19. While we did not declare in this paper that we have solve all the problems in epidemiological modelling, however on a serious note we can say that by introduction of an indicator function, we were able to capture variabilities of the spread more importantly, we were able to predict waves. Using several differential operators we were able to capture not only the daily numbers of new infected.

## Data Availability

World Health Organization, WHO Coronavirus Disease (Covid-19) Dashboard, https://covid19.who.int/table.

## Notes

### Competing Interest Statement

The authors have declared no competing interest.

### Funding Statement

This study did not receive any funding

